# Quaternary Prevention: Is this Concept Relevant to Public Health? A Bibliometric and Descriptive Content Analysis

**DOI:** 10.1101/19007526

**Authors:** Miguel Andino Depallens, Jane Mary de Medeiros Guimarães, Naomar Almeida Filho

## Abstract

**Objective:** To measure and map research output on Quaternary Prevention (P4) and outline research trends; to assess the papers content, mainly regarding methods and subjects approached in order to contribute to the improvement of global knowledge about P4 and to evaluate its relevance for public health.

**Design:** Bibliometric and descriptive content analysis.

**Articles reviewed:** Scientific articles about P4 recorded in Pubmed, LILACS, Scielo or CINAHL published until August 2018, with correspondent full articles available in Portuguese, English, Spanish, German or French.

**Main outcome measures:** Year of publication, first authors’ name and nationality, journals’ name, country and ranking, publication language, used methods and main reported subjects.

**Results:** 65 articles were included, published in 33 journals of 16 countries between 2003 and 2018 with a peak of publications in 2015. The first authors came from 17 different countries, 23% of them were Brazilian and Uruguay was the leading nation according to the scientific production per capita. 40% of all the selected articles were in English, 32% in Portuguese, 26% in Spanish. 28% of the papers were published in Q1 or Q2 journals. The research outputs on P4 begun first in the South of Europe, went to South America and then expanded worldwide. 88% of the articles were bibliographic research and 38% of all focused on specific examples of medical overuse (including several screening tests).

**Conclusions:** Quaternary prevention represents an ethical and valid approach to prevent occurence of iatrogenic events and to achieve equal and fair access to health services. Conceptual, geographical and linguistic elements, as well as WONCA conferences and type of healthcare systems in the authors’ country were fundamental factors that affected research output. The quality and quantity of available studies is still limited, therefore further investigations are recommended to assess the effective impact of P4 on public health.

## Introduction

The general notion of prevention has been instrumental for health care provision since early years in Ancient Greece. The simple idea that human beings should be protected from the influence of factors potentially harmful to their health indeed guided modern forms of medical practice (clinical, pharmacological and surgical interventions) and public health (sanitation, vaccination etc.) oriented by scientific criteria. These developments took place initially in Western Europe, during the Enlightenment era, resulting in the movements of Hygiene, Medical Police and Social Medicine.^1^ The institutionalization and spread of preventive practices and measures in Medicine and Public Health were consolidated during the first half of the XXth century, largely due to Leavell & Clark’s ternary model of types or levels of prevention: primary, secondary and tertiary.^2^

The idea that medical practice in all manifestations could be fully described as different modalities or levels of preventive action have only prevailed particularly since World War II. However, despite strong efforts towards integrality and equity in health care, such as the WHO’s Health for All campaign, only late in past century criticisms and conceptual advances were brought about concerning this subject-matter.^3^ Among them, a definition of “quaternary prevention” regarding iatrogenic effects, as proposed the Belgian family physician Marc Jamoulle (1986)^4^, was an add-on to the theory of levels of prevention. This novel conceptual and practical approach may have enormous potential to foster better-quality, socially referenced, more humane health care systems and practices, needed especially in these days of widespread budget cuts and exclusionary policies in public health services, all over the world.

The main objective of this paper is to report a bibliometric and descriptive content analysis of research output on the concept of Quaternary Prevention (P4) published until August of 2018, and to prospectively correlate research trends. To achieve such goals, first, we briefly provide some theoretical and methodological background, and context information on the historical development of prevention approaches. Second, we present methods, describe the criteria for considering the papers and point the types of data that were collected, analyzed and classified, in order to measure and map scientific production as well as to assess the papers content. Third, we expose the results, focusing on: year of publication, authors’ and journals’ features, methods used in the selected studies and main topics approached. And fourth, we outline and discuss some trends towards research on Quaternary Prevention, emphasizing the main methods and subjects treated.

We hope to contribute to improve the global knowledge of P4 in order to further the understanding of this recent and relevant concept, highlighting authors, countries and journals involved in the research and publications on this topic and to classify the selected articles. In further papers, we intend to report a critical synthesis and in-depth analyses of the selected articles in a qualitative systematic review.

### Quaternary Prevention: Conceptual Background

The term iatrogenesis comes from the Greek language: “iatro” (medicine) and “genesis” (creation); it can be defined as medical harm or adverse effect. Iatrogenic events always existed in historical times. The Hippocratic oath (450-350 B. C. E.) confirms this statement and the awareness of the Ancient Greece physicians of the potential iatrogenic outcomes that could result from their practice. Across the centuries, the beneficence and non-maleficence principles – *primum non nocere* (first do no harm) – became one of the cornerstones of current bioethics.^5^

Over the course of the next 20 centuries, there was little increase in the effectiveness of healing therapies, remained not much effective. However, in those times, a few researchers produced very relevant public health knowledge and tangible strategies became available in the field of disease prevention. In this perspective, James Lind (1716–1794) established that the ingestion of some specific nutrients, like vitamin C, could prevent diseases, scurvy in this case. In a corresponding period, public health and prevention of disease became a priority for a few bellicose states that needed strong soldiers and workers. Johan Peter Frank (1745-1821) in Germany, implemented massive health policies to tackle infectious outbreaks related to hospitals, military medicine, litter sanitation, food and housing. Then, Jenner (1749-1823), Pasteur (1822-1895) and Koch (1843-1910) discovered the pathogenic role of some microorganisms and developed effective vaccines to prevent the related diseases (e. g.: smallpox, rabies). And the basics of modern epidemiology were established during the cholera outbreak in London (1849) by John Snow (1813–1858), who discovered the pathogenic role of infected water, mapped the cases of cholera and identified the contaminated spread of the sources, enabling the prevention of its dissemination.^6^

Spectacular individual and biochemical treatments followed upon the discovery of insulin therapy, by Banting (1922) and his team that enabled to save young diabetic end-stage patients from certain death.^7^ A few years later, in 1928, Fleming (1881-1955) made another fabulous finding and discovered the penicillin that open the period of curative therapies. Around 1940, Sir Archibald Cochrane (1909-1988) developed a new and very significant concept: the evidence-based medicine (EBM). In this way, the randomized clinical trial (RCT) came to be considered a revolutionary scientific method, which enable the comparison of two different therapies.^8^ Up to the present, RCTs are used worldwide to guide most of clinical decision-making.

At the end of the 1980s, influenced by Illich’s book about iatrogenesis^9^, the Belgian family physician Marc Jamoulle revised^10^ the prevention levels of Leavell and Clark^11^, integrating a public health approach into individual clinical practice. His classification was designed according to two specific dimensions: one defined by the patient’s feeling (illness) and the other by the clinical assessment (disease)^12^. We put hereinafter the definition of the levels of prevention together with the initial approach of Jamoulle:

1. Primary prevention (P1) acts on healthy individual (absence of illness) to avoid the incidence of a specific disease (absence of disease), for example, through counseling about smoking cessation to prevent lung cancer and chronic obstructive pulmonary disease (COPD);
2. Secondary prevention (P2) aims to detect early a potentially severe disease in an asymptomatic population (absence of illness), increasing the probability of cure by identifying and treating an initial stage of the detected disease (presence of disease), for example through a screening method (e. g.: annual eye funduscopy to detect diabetic retinopathy).
3. Tertiary prevention (P3) proposes to reduce the impacts of any disease on the quality of life (illness and disease present) through medical treatment or rehabilitation, for example through an earlier neurorehabilitation therapy after a disabling stroke.
4. Quaternary prevention (P4, first definition) aims to prevent medical overuse in situations where the patient feels ill (illness present), but the physician does not link the symptoms to any biological disease (absence of disease). An example of P4 could be the use of a “watchful waiting” strategy (watch and follow-up) when a young healthy patient without any cardiovascular risk or symptom worries about his cholesterol level.

We can define overmedicalization or medical overuse as a medical intervention that brings more harm than benefit.^13^ In this situation and if a drug was prescribed to the young patient, it would be considered a typical case of overmedicalization. There are plenty of medical overuse/P4 situations and we will see some in further topics.

In 2000, Barbara Starfield reported data showing that iatrogenic events represented the third cause of death in the United States.^15^ Moreover, several authors were researching medical overuse and P4, denouncing and attempting to prevent iatrogenic procedures.^16, 17, 18, 19^ In 2003, the concept of P4 was finally acknowledged by the scientific community and the World Organization of National Colleges, Academies and Academic Associations of General Practitioners/Family Physicians (WONCA) International Classification Committee. The following definition has been accepted: “Action taken to identify a patient or a population at risk of overmedicalization, to protect them from new medical invasions and suggest to them interventions, which are ethically acceptable”.^20^ This new definition has an enhanced scope and is supported by the nonmaleficence bioethical principle. It aims to prevent all kinds of medical overuse without considering the features of the clinical situation, neither the patient’s feeling nor the physician’s assessment. Figure 1 may help in understanding both the initial and actual definition, and how P4 can be applied according to the specificities of some clinical situations.

**Figure 1.**
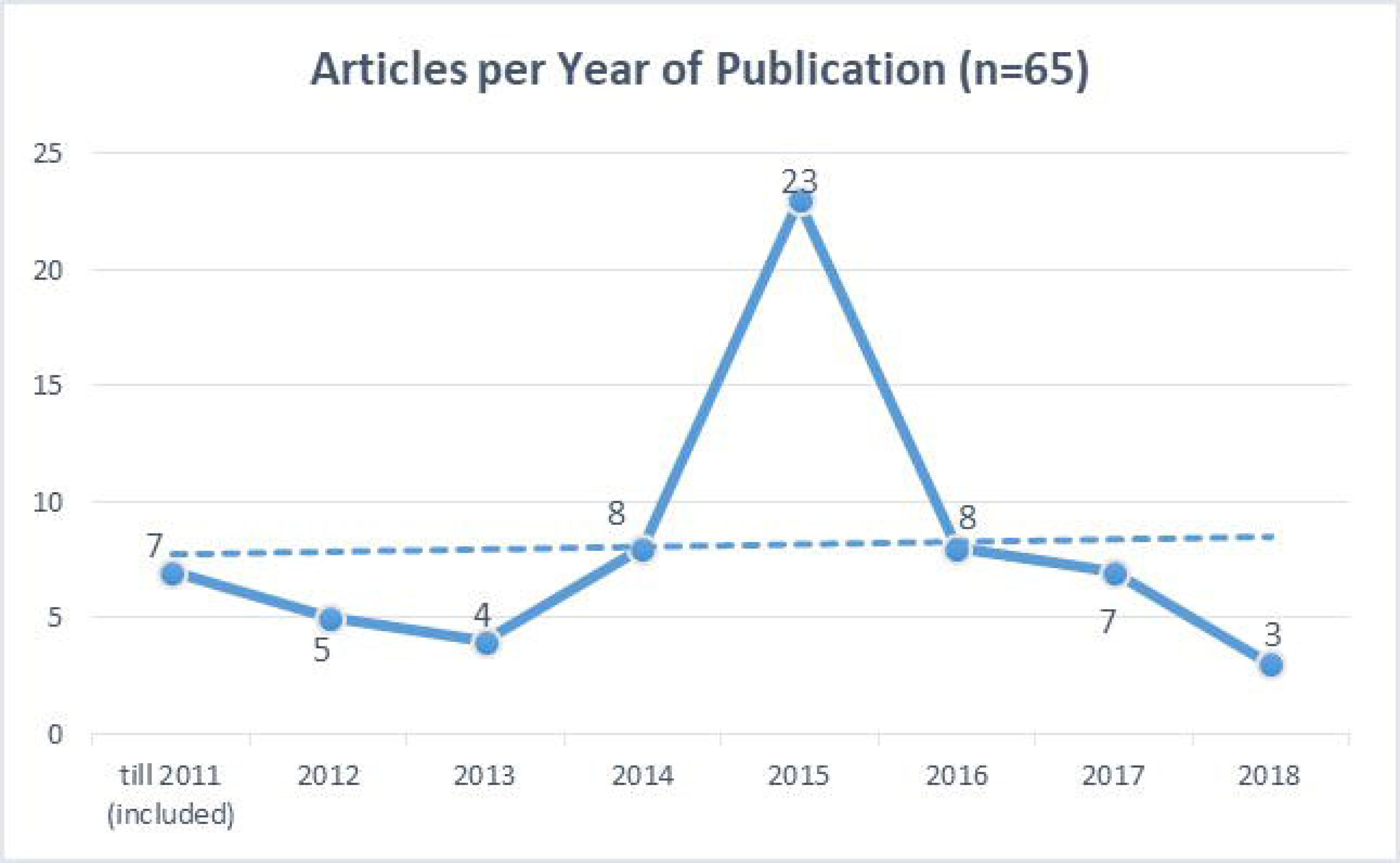
Quaternary Prevention in all the levels of prevention according to the patient’s feeling and the physician’s assessment (adapted from Jamoulle^14^).

As well as the prevention of medical overuse and iatrogenesis, P4 also aims to significantly lower some related specific costs of overall health expenditure responding to one of the actual worldwide governments’ main objectives.^21, 22^ Consequently, P4 could allow the expansion of global health coverage and confirms its relevance for public health.

## Methods

To achieve an integral assessment of research output on P4, two complementary methods were used: a quantitative bibliometric approach and a qualitative content analysis. Bibliometrics is a methodology which focuses on measuring and evaluating research trends.^23, 24^ Topics of bibliometric studies on adjacent topics are: medicalization during pregnancy,^25^ overdiagnosis,^26^ which emphasized breast cancer screening. Also, we found two more papers about “unnecessary procedures”.^27, 28^ However, these studies focused on specific examples of overmedicalization or had significantly different objectives (e.g.: focusing the impact of Cochrane Reviews on clinical decision); none of them approached P4. The criteria of the Cochrane Handbook^29^ have been used to build the search strategy and the papers’ selection process. “Evidence from qualitative studies can play an important role in adding value to systematic reviews for policy, practice and consumer decision-making”,^30^ therefore, complementing the bibliometric quantitative approach, a content analysis has been achieved according to the Standards for Reporting Qualitative Research (SRQR)^31^ and constitutes the second part of the present article.

### Criteria for considering articles for this review

We included all scientific articles, published up until August, 7^th^, 2018, from the research databases, available in Portuguese, English, Spanish, German or French, and using the WICC (2003) definition’s meaning when it mentioned P4. We excluded articles that were commentary, letter or editorial as well as documents that were not articles, such as: master or doctorate thesis, teaching material, videos, among others, as well as all other records that did not fit our inclusion criteria.

### Data and outcomes

The following data were collected: title; name of the author(s) and their nationality; year of publication; name of the journal and country of its headquarters; publication’s language; methods; abstracts.

### Search methods for identification of articles

We did the search using the following databases: Pubmed, LILACS, Scielo and CINAHL. There were no existing records about P4 in the Cochrane Library; for this reason, this renowned database was not included. P4 is a relatively recent concept and thus there is not much literature about it, therefore we decided to use the widest possible Search Builder: using the conjunction “OR” and, when available, coupled with a Medical Subject Headings (MeSH). In Pubmed, Scielo and CINAHL, there was no pre-existing MeSH about Quaternary Prevention; therefore, we used the following Search Builder: “Quaternary Prevention” OR “Prevención Cuaternaria” OR “Prevenção Quaternária“. In LILACS, there was already a descriptor (*Descritores em Ciências da Saúde*: DECS) - “Quaternary Prevention” - that corresponds to a MeSH in the Pubmed database. So, we expanded our Search Builder in LILACS as follows: (tw:(“prevenção quaternária” OR “quaternary prevention” OR “prevención cuaternária”)) OR (mh:(prevenção quaternária)). To illustrate the selection process of the papers, we used Review Manager 5.3**®** and designed a flow diagram (figure 2).

**Figure 2.**
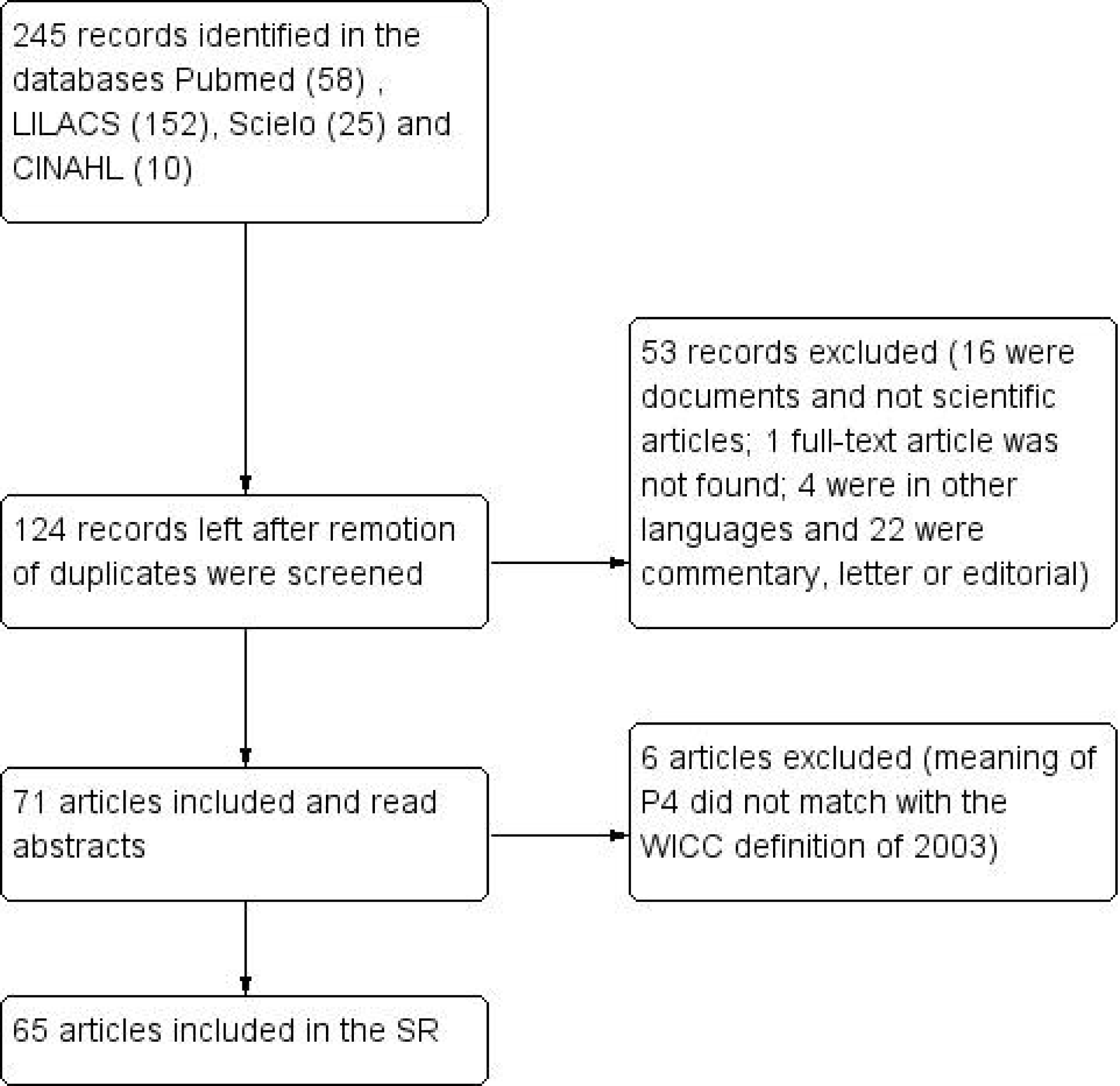
Flow chart of the articles selection.

### Data collection, analysis and extraction

The searches were exported directly from the databases to Zotero**®**, which accelerated the transfer of information, the removal of duplicates and facilitated the achievement of the initial bibliographic list with a specific tool. Results from electronic databases searches were reviewed to check the inclusion and exclusion criteria. The abstracts were read, the papers classified and the extracted was digitalized in Excel**®** tables. The articles that were not freely available have been accessed through the CAPES (*Coordenação de Aperfeiçoamento de Pessoal de Nível Superior*) website.^32^ We read the abstracts of the selected articles to check the meaning of P4 and, if necessary, excluded the ones which used a very different definition. Finally, we searched for the journal rank in two acknowledged databases: one European – Scimago^33^ – and one South American institution – Qualis/CAPES.^34^ The journal ranks were entered into the Excel**®** tables, together with previously collected data.

### Bibliometric analysis

We conducted the bibliometric analysis, building dynamic cross tables and outlining some trends according to the following data: year of publication; language of publication; name of the first authors and nationality; Publications per capita (Number of publications per first author’s country / number of residents in million); The data of each country’s population in 2019 has been obtained calculating an estimation stem from the *United Nations population estimates and projections*^35^; name of the journal and country of its headquarters; journal ranking (2017 Scimago: quartile Q1-Q5); journal ranking (2013-2016 Qualis/CAPES: A1-C).

### Content analysis

Focusing on the title and abstract, we categorized each paper according to its method, such as: bibliographic research, qualitative research, quantitative research, case report, among others. Articles were also classified under one of the following categories:

1. Epistemological and conceptual research focusing on P4 general concept and/or the overmedicalization process;
2. Studies addressing bioethical issues related to P4, discussing in particular the non-maleficence and the justice principles;
3. Reviews that consider empirical evidence on medical overuse and iatrogenesis, highlighting examples of overmedicalization (except screening);
4. Screening also represents a type of potential iatrogenesis, so we separated it from the previous category because it is one of the main focus of P4;
5. Investigations that outline P4 implementation strategies and/or aim to divulge the concept.

We identified 245 records in the electronic databases: 152 in LILACS, 58 in Pubmed, 25 in Scielo and 10 in CINAHL. 121 duplicates were removed, and the remaining 124 records left were screened. We excluded one record, because we did not find it even after sending an email to the journal and the author (Gofrit, 2000); 16 because they were not scientific articles (master or doctorate thesis, teaching material, MeSH, second profissional opinion or videos), four full-articles were in other languages (three in Serbo-Croat and one in Corean) and 22 were commentaries, letters or editorials. After these steps, we read the 71 abstracts left and excluded 6 more articles (Gadelha, 1992; McColl and al., 2017; Moses, Mawby, Phillips, 2013; Trova and al., 2015; Weinstein, 2001) as they used a different definition of P4, defining it like a physical or social rehabilitation, which did not match with the meaning of the definition accepted by the WICC (2003). Finally, 65 articles were included in the study (Figure 2).

## Results

### Bibliometric analysis

The 65 selected articles were published between 2003 and 2018. Until 2011, there was less than one publication a year. Starting from 2012, we observe a slow increase in the scientific output untill 2014; 2015 represents a peak with 23 publications (35% of the total number). After 2015 (2016-2018), the publications returns to their basal numbers, similar to the 2012-2014 period (Figure 3).

**Figure 3.**
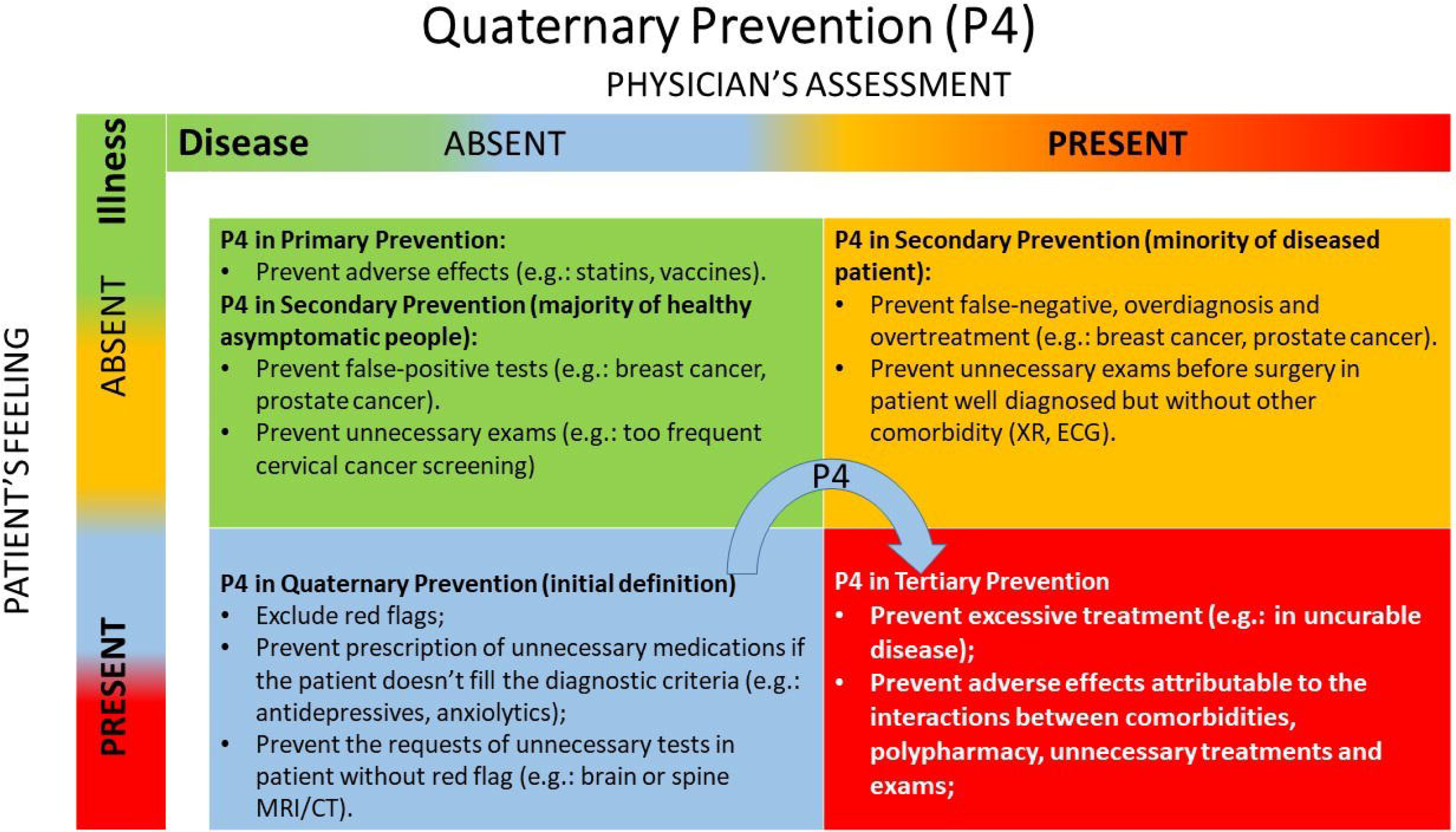
Number of papers according to the year of publication (n=65).

#### Authors

The first authors came from 17 different countries in Europe, South and North America and Asia. Brazilian authors were the leaders of 15 papers (23%), the Spanish were in the second position with 11 published articles (17%) followed by the Portuguese with 8 publications (12%) (Table 1). To consider the population size of each countries, we calculated the scientific production per capita. Consequently, we reached another ranking in favour of the smaller countries: Uruguay was first (1,18), Portugal (0,80) moved up to the second place and there were three countries with a very close index in the third place: Belgium (0,36), Switzerland (0,24) and Spain (0,23); Brazil (0,07) fell to the 7^th^ position.

**Table 1.** Number of articles according to first author’s nationality and year of publication (n=65) and production per capita.

|  | till 2011<br>(included) | 2012 | 2013 | 2014 | 2015 | 2016 | 2017 | 2018 | Total | Prod.<br>p/capita (in<br>millions) |
| --- | --- | --- | --- | --- | --- | --- | --- | --- | --- | --- |
| Brazil | 1 | 2 |  | 4 | 3 | 2 | 2 | 1 | 15 | 0,07 |
| Spain | 6 | 1 | 1 | 1 | 2 |  |  |  | 11 | 0,23 |
| Portugal |  | 2 | 1 | 1 | 3 |  |  | 1 | 8 | 0,80 |
| Argentina |  |  |  |  | 3 |  | 2 |  | 5 | 0,11 |
| Belgium |  |  |  | 1 | 3 |  |  |  | 4 | 0,36 |
| Uruguay |  |  |  |  | 2 | 1 | 1 |  | 4 | 1,18 |
| India |  |  |  |  | 1 | 2 |  |  | 3 | 0,00 |
| France |  |  |  |  | 1 |  |  | 1 | 2 | 0,03 |
| South Korea |  |  |  |  | 1 | 1 |  |  | 2 | 0,04 |
| Switzerland |  |  |  | 1 | 1 |  |  |  | 2 | 0,24 |
| Peru |  |  |  |  |  | 1 | 1 |  | 2 | 0,06 |
| Canada |  |  |  |  | 2 |  |  |  | 2 | 0,05 |
| Germany |  |  |  |  |  |  | 1 |  | 1 | 0,01 |
| Vietnam |  |  |  |  | 1 |  |  |  | 1 | 0,01 |
| Norway |  |  |  |  |  | 1 |  |  | 1 | 0,2 |
| Bolivia |  |  | 1 |  |  |  |  |  | 1 | 0,09 |
| Italy |  |  | 1 |  |  |  |  |  | 1 | 0,02 |
| Total | 7 | 5 | 4 | 8 | 23 | 8 | 7 | 3 | 65 | 0,22 |

Until 2011, six out of seven articles were written by Spanish authors and, in particular, three by Juán Gérvas (2003, 2006, 2006) - he wrote another article in 2012. Gérvas is a family physician. He is a retired professor from the University of Valladolid (Spain) and the Johns Hopkins University (United States), committed researcher who published more than 400 articles in scientific journals, and has been a member of the WICC since 1986. He is one of the most active proponents of P4 and wrote one of the most significant books about this topic, *Sano y salvo* (“Healthy and Safe”, 2012). Then, Norman and Tesser, promoted the concept in South America through their first article about P4 (2009). Both are Brazilian family physician, professors and researchers. We noted that untill 2012, there were only Spanish, Brazilian and Portuguese authors who wrote about P4. Tesser, professor of the Federal University of Santa Catarina and of the University of Coimbra, and Jamoulle, the “father” of P4, family physician and researcher, are the most established authors on the subject of P4. Both had six articles included here. Beyond Europe and South America, 11 out of 17 (65%) authors came from Latin-speaking countries, we can see that Asian authors from South Korea, India and Vietnam, as much as North America, with Canadian authors also took part in this process map 1).

**Map 1.**
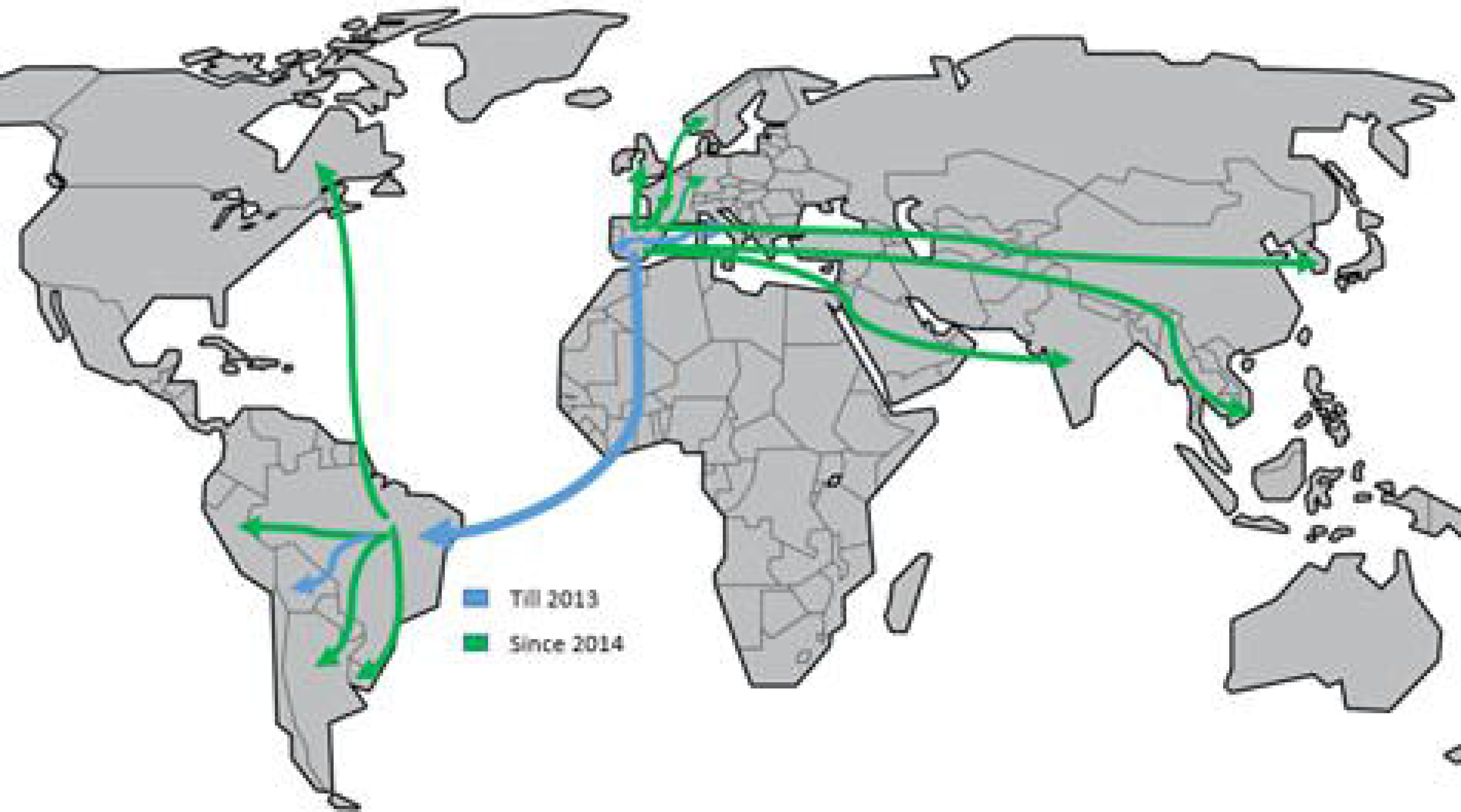
Map of the expansion of the publications about P4 according to first author’s nationality.

#### Languages

The English language has been used in 40% of the papers, 32% were in Portuguese, 26% in Spanish. There was only one publication available in French (1.5%) published by Widmer (Switzerland) and Jamoulle (Belgium) in the Revue Médicale Suisse, the main Swiss medical journal. So we can conclude that 60% of the articles were written in a Latin-spoken language. Until 2011, the papers were written mainly by Spanish authors like Gérvas, in Spanish language and none of these were published in English (Table 2 or Figure 4). Between 2012 and 2014, 88% of the papers were in Spanish or Portuguese. In 2015, a significant increase in papers written in English was seen, 14 exactly (60% of the articles published the same year) written by European, Asian and South American authors. After 2015, the languages of publication became more balanced between English, Spanish and Portuguese.

**Table 2.** Number of publications according to the year and language of publication (n=65)

|  | till 2011<br>(included) | 2012 | 2013 | 2014 | 2015 | 2016 | 2017 | 2018 | Total |
| --- | --- | --- | --- | --- | --- | --- | --- | --- | --- |
| English |  | 1 | 1 |  | 14 | 5 | 3 | 2 | 26 |
| Spanish | 6 | 1 | 2 | 1 | 5 | 2 | 4 |  | 21 |
| Portuguese | 1 | 3 | 1 | 6 | 4 | 1 |  | 1 | 17 |
| French |  |  |  | 1 |  |  |  |  | 1 |
| **Total** | **7** | **5** | **4** | **8** | **23** | **8** | **7** | **3** | **65** |

#### Journals

The table 3 shows that 33 journals of 16 countries published articles on P4 worldwide. The chronological and geographical expansion of the P4 publications according to the first author’s nationality was quite comparable to that of the journal’s headquarter countries. The first published articles started in the South of Europe, then spread to Brazil and reached afterward North America and Asia. There was a clear predominance of 9 Brazilian journals that published 33 papers (51% of all articles). The authors, generally, published in journals of their own country or of a close region. We can build some associations between the author’s nationality and the journals country, such as: Brazilian authors published 14/15 papers in Brazilian journals; Spanish authors published 9/11 articles in Spanish journals; Portuguese authors published 5/8 studies in Brazilian Journals; Argentinian authors published 4/5 papers in Argentinian journals; Uruguayan published 3/4 in Brazilian journals and Indian authors published 3/3 articles in a Pakistani journal. Furthermore, as we wrote above, a significant majority of authors published in Brazilian journals.

**Table 3.**
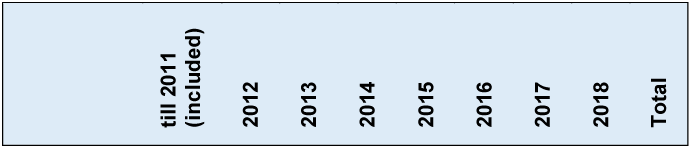
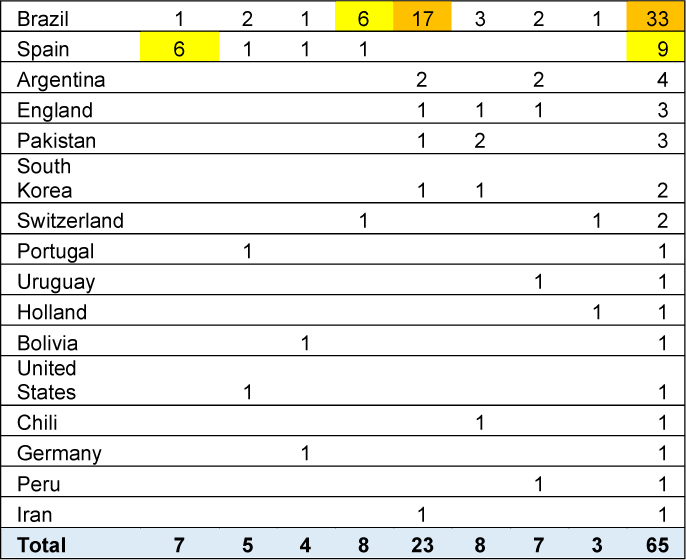
Number of articles according to the journal headquarter’s country and the year of publication (n=65)

| till 2011<br>(included) | 2012 | 2013 | 2014 | 2015 | 2016 | 2017 | 2018 | Total |
| --- | --- | --- | --- | --- | --- | --- | --- | --- |

|  |  |  |  |  |  |  |  |  |  |
| --- | --- | --- | --- | --- | --- | --- | --- | --- | --- |
| Brazil | 1 | 2 | 1 | 6 | 17 | 3 | 2 | 1 | 33 |
| Spain | 6 | 1 | 1 | 1 |  |  |  |  | 9 |
| Argentina |  |  |  |  | 2 |  | 2 |  | 4 |
| England |  |  |  |  | 1 | 1 | 1 |  | 3 |
| Pakistan |  |  |  |  | 1 | 2 |  |  | 3 |
| South Korea |  |  |  |  | 1 | 1 |  |  | 2 |
| Switzerland |  |  |  | 1 |  |  |  | 1 | 2 |
| Portugal |  | 1 |  |  |  |  |  |  | 1 |
| Uruguay |  |  |  |  |  |  | 1 |  | 1 |
| Holland |  |  |  |  |  |  |  | 1 | 1 |
| Bolivia |  |  | 1 |  |  |  |  |  | 1 |
| United States |  | 1 |  |  |  |  |  |  | 1 |
| Chili |  |  |  |  |  | 1 |  |  | 1 |
| Germany |  |  | 1 |  |  |  |  |  | 1 |
| Peru |  |  |  |  |  |  | 1 |  | 1 |
| Iran |  |  |  |  | 1 |  |  |  | 1 |
| Total | 7 | 5 | 4 | 8 | 23 | 8 | 7 | 3 | 65 |

The Brazilian Journal of Family and Community Medicine (RBMFC) had the highest number of accepted articles about P4. They published 22 papers (34% of all) between 2013 and 2016, 10 articles were in English (all in 2015), 8 in Portuguese, 4 in Spanish and attracted multiple authors of different nationalities (Argentina, Belgium, Brazil, Canada, France, Portugal, Spain, Switzerland, Uruguay and Vietnam).

According to Table 4, we see that the majority of papers (49% Scimago; 37% Qualis/CAPES) were published in journals not included in the international rankings, neither in the Scimago (Europe), nor in the Qualis/CAPES (Brazil). 28% were published in Q1 or Q2 journals. Only French, German, Italian and Portuguese authors achieved a publication in the following Q1 journals, all were published in English, respectively in: Monographs in Oral Science (Switzerland), BMC family practice (England), Archives of Gynecology and Obstetrics (Germany) and SpringerPlus (England). The main Q2 journals were: Gaceta Sanitaria (Spain; 3 articles by Spanish first authors; all in Spanish), Cadernos de Saúde Pública (Brazil; 3 articles by Brazilian first authors; 2 in Portuguese and 1 in English) and Journal of Preventive Medicine and Public Health (South Corea; 2 articles by a South Corean first author; 2 papers in English). We also observed that the publications in Q1 and Q2 journals are stable or slowly increasing, however the publication in non-classified journals are decreasing significantly.

**Table 4.**
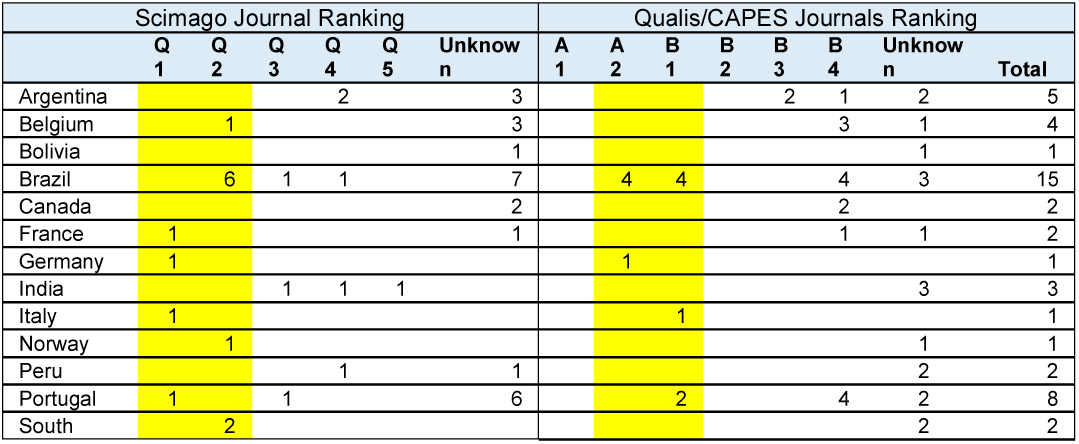
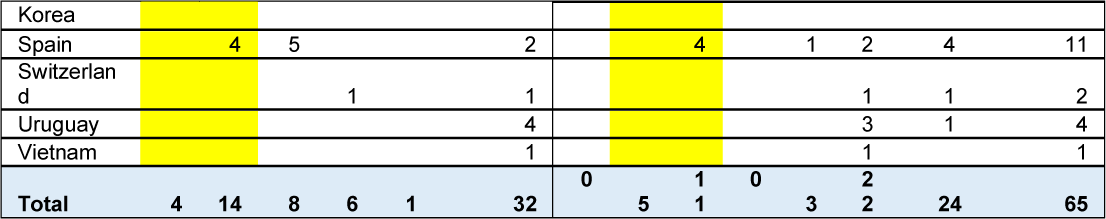
Number of articles according to the journal rank (Scimago Quartile and Qualis/CAPES) and first author’s nationality.

#### Methods of the selected papers

The vast majority of the articles (88%) used a bibliographic research as their main method. There were also 5% of papers that used case reports and the same ratio used qualitative research (e. g. interviewing family physician about P4); quantitative research was quite rare with only one article (1%).

#### Main categories

According to the distribution of the main approached topics, 16 articles (25%) were graded in the conceptual category “general considerations about P4 and overmedicalization process”. We classified many of the discussed aspects as: theoretical and conceptual aspects (levels of prevention, EBM, distinction between preventive approach and clinical care), medicalization drivers and consequences (conflict of interest, disease mongering, medicalization of risk factors, overdiagnosis and overtreatment) and protective factors (PHC attributes: longitudinality and role of gate-keeper; communication skills: doctor-patient relationship, patient-based medicine; Bioethics: non-maleficence and justice principles; and patient-safety strategies.

Only four articles (6%) were graded in “Bioethics”, approaching many topics, such as: the definition of health; the humanization of the relationship doctor-patient; the clinical context of uncertainty; political, economic and social determinants in health; autonomy, beneficence, non-maleficence and justice principles; intellectual freedom and responsibility; epistemology of medicine.

We classified 32 papers (49% of all) under the category “specific examples of overmedicalization and iatrogenesis”, including “screening”. 31% belong to “specific iatrogenic situations”, like chronic diseases and polypharmacy (e. g.: diabetes, genetic syndrome), birth, mental health and others (odontology, geriatrics, pathology) and 18% belong to the “screening” category (general aspects, cervical, breast and prostate cancers, genetic testing and neonatal screening).

And 13 articles (20%) were categorized as belonging to the subject “implementation and divulgation strategies of P4”: four of them developed the humanization of health assistance and the major role of the communication skills, such as the patient-centered medicine, to reduce iatrogenic events. The other articles discussed medical prescription, strategies to ensure patient’s safety, applicability of “Choosing Wisely” campaigns, manifestoes in favour of P4 and defending a medicine without conflict of interests, P4 teaching and search for P4 in electronic databases.

## Discussion

To understand the timeline of research output on P4, we need to remember the substancial publications of 2000 and 2002 about iatrogenesis, such as: “Is US health really the best in the world?”,^36^ “The arrogance of preventive medicine”^37^ and “Too much medicine?”.^38^ Moreover, Gérvas had already published a paper in this respect in 1997, reporting the inefficiency of statin treatment in primary prevention of ischemic cardiopathy.^39^ In this this context, the WICC accepted the definition of P4 in 2003, acknowledging its relevance and encouraging future publications. It is exactly the same year that Gérvas published the first article included in our review.^40^ Afterwards, the scientific output increased slowly and progressively until its peak in 2015 which was probably linked to the disclosure, in February 2015: P4 would be one of the main subjects of the 21^st^ WONCA (World Organization of Family Doctors) World Conference 2016 in Rio de Janeiro. Consequently, this reputed event encouraged many international authors to undergo research and publish on the subject of P4 (figure 3). Furthermore, the same year, Jamoulle led a specific edition of the RBMFC about P4 and the Iberoamerican Family and Community Medicine Conference took place in Uruguay, also emphasized quaternary prevention. Those events led to a significant increase in the publications in English, together with a geographical expansion of the first authors countries. After 2015, the scientific output declined and came back to its baseline.

Until 2012, the first authors came from Spain, Portugal and Brazil, countries that have universal public health system. These systems, by definition, are very concerned about equity, which is one of the P4 main objectives; it can be achieved totally or partially, through the reduction of unnecessary medical procedures, which, indirectly, promotes equity and favors the access to health services. We also demonstrated that Latin-speaking countries first authors seem to be more acquainted with the concept and published more about it; therefore the same countries occupied the three first positions of the publications ranking. However, when we assessed the number of papers per capita, Uruguay was first. It is a very small country of 3,47 million people that lead the WONCA group on P4, which also has a universal public health system and borders the South of Brazil and is greatly influenced by its larger neighbour of 212 million of inhabitants.^41^

The results reported a clear predominance of Brazilian journals and we suggest at least six reasons to explain it. Firstly, the Brazilian authors, Norman and Tesser,^42^ were precursors of the concept in Latin America since 2009, which promoted the submission of articles in Brazilian journals. Secondly, because Brazil is one of the biggest universal healthcare systems in the world and suffer from insufficient funding, It requires a cost-effective management that can be achieved, at least partly, with P4 implementation policies, tackling medical overuse. Thirdly, because Brazilian journals accept articles in many languages, drawing international authors. Fourthly, as we wrote above, P4 was one of the main subjects in the World WONCA Conference of Rio de Janeiro in 2016, and it encouraged the Brazilian journals and authors to publish on this subject. Fifthly, the municipality of Rio de Janeiro, reformed completely and enhanced its primary health care (PHC) system in 2009-2012,^43^ improving the access of the population to health services, emphasizing universality and equity principles. And sixthly, the Brazilian government launched a massive program in 2013 to strengthen its national PHC, the “More Doctors Program” (*Programa Mais Médicos*) based on the principles of universality and equity, engaging more than 18.000 national and foreign physicians, favouring the access of 63 million of Brazilians to the health system.^44^

Most of the papers were published in non-classified journals. However, a significant number (18 articles, 28%) also published in Q1 or Q2 journals. Some readers could deduct that the majority of the papers were published in low quality journals. However, according to Seglen (1997):

> - Journals’ impact factors are determined by technicalities unrelated to the scientific quality of their articles;
> - Journal impact factors depend on the research field: high impact factors are likely in journals covering large areas of basic research with a rapidly expanding but short lived literature that use many references per article.^45^

In this regard, some determinant factors, like the language of publication, can strongly impact the number of citations and, consequently, affect the journal impact factor and the article’s visibility. Therefore, if P4 is published in non-classified journals, it can be one more factor that impairs its disclosure and, in this way, its expansion. Another aspect can possibly explain the publications in low impact factor journals: publication costs. Thus, most of the selected studies have been realized in low or middle-income countries, without any sponsorship and probably could not afford high cost of publication in high impact factor journals. Nevertheless, according to the four publications in Q1 journals, we note four characteristics that could have promoted them in the best classified journals. First, the geographical situation promoted the publications of European authors in European Journals. Second, all of them used the English language that constitutes the main official language of the European Union. Third, we also observed that three of the four first authors were from Latin-speaking countries (France, Italy and Portugal) that confirmed the approximation between such countries and the concept of P4. And fourth, the economic aspect could have facilitated authors from high-income countries to pay the expensive publication costs of high impact journals.

Assessing the quality of the selected papers has been a quite difficult and sensitive task, especially because most of the articles were literature reviews and that type of study rarely uses a structured methodology; we confirm this trend in our study. In addition, while the papers frequently brought a broad conceptual understanding, including multidimensional analysis, they usually supported P4 thesis and seldom critically developed the potential weaknesses of the concept.

P4 is a complex concept based on historical, theoretical and scientific fundamentals that involves several areas of knowledge, suggesting the necessity of an exhaustive expertise in these areas (e. g.: politics, economics, sociology, law, public health management, history and epistemology of medicine, medical teaching, professional practice, EBM, bioethics, psychology, communication skills, among others). The previous understanding of these basics and their interconnections is essential and enable the development of effective interventions to tackle unnecessary medical procedures and their iatrogenic effects. Therefore, the category “general considerations about P4 and overmedicalization process” is more than necessary to be developed. The overmedicalization process is described as an effect of biased studies, conflict of interest and disease mongering strategies, involving mainly the pharmaceutical industry but also some physicians.^46^ In this regard, considerable advertising campaigns enhanced this phenomenon; it lead the population to a more concerned condition about their own health and raised the expectations according to the health care system, even when healthy. Concerning the definition of P4, the authors wrote it generally just after the first three levels of prevention of Leavell and Clark.

Quaternary prevention constitutes a counter-hegemonic concept, which approaches modern and occidental medicine being based on a critical and multidimensional perspective. Moreover, actual biomedical research, regardless of its funding,^47, 48^ focuses on high-technology studies^49^ and does not prioritize medical overuse issues. Therefore, we understand why the vast majority of the included papers were bibliographic research and low cost studies. Bibliographic studies were also necessary to create the robust theoretic base of such a recent and interdisciplinary concept. Qualitative studies are essential to promote the identification and analysis of key elements involved in the medical overuse process, for example regarding the perception and practice of the health actors. In this way, these points are precious because they can lead to the elaboration of effective strategies, in order to achieve significant changes in health practice and impact medical overuse. However, only a small number of qualitative studies were found, probably because they were very time-consuming and of high cost. Quantitative research was also very scarce, probably because it is generally used to evidence the efficacy of a medical intervention and rarely to establish its harm. However, the only quantitative included study^50^ highlighted significant data about overscreening and underscreening for cervical cancer in a specific population. This type of study is brief, low-cost and very relevant, because able to reveal precise quantitative estimations about overmedicalization procedures, including the expenditure that can be saved if a P4 approach would be implemented. Case-report studies can also be used to improve theoretical aspects on P4 but have less potential to contribute to elaborate concrete strategy to impact overmedicalization because they stem from the analysis of a singular and practical example.

In a context where medicine is inducing much medical overuse, leading to an outbreak of iatrogenic events^51^ and is, by definition, contrary to the principles of medicine; the papers approached this issue through an ethical perspective. This point represents the heart of the P4 problematic and implies to consider the risk and benefits of each clinical decision, integrating a constant critical reflection on beneficence and non-maleficence principle into health practice. A broad theoretical knowledge, accurate competence to assess critically the available evidences and a global understanding of each clinical case are required to apply such type of practice. The second point focuses on the autonomy principle, since the patient always takes part in the clinical decision-making. In some situations, its role can be more significant, especially in case of shared decision. Besides, plenty of other factors are also able to affect the patient’s opinion, for example: an advertisement which minimizes the risks of a medical intervention (e. g. PSA screening^52^) and does not mention the prevalence of severe iatrogenic damages. This situation illustrates well a possibility of bias in the free and informed patient decision-making concerning a single screening test. In this case, the role of P4 is to identify such situations and be able to build strategies to face or prevent them.

Last but not least, P4 aims to tackle medical overuse, saving funding and allowing some necessary medical interventions which could not be realized because of insufficient funding. This last example highlights the role of P4 advocating the justice principle. In this category, the authors considered some conceptual aspects as: knowledge (epistemology) and ethics (non-maleficence and beneficence principles); analyzed specific bioethical challenges in the case of “incidentalomas”;^53^ outlined schemes, remembering the importance of cross-sectoral engagement, including political, economic and social dimensions, to recover the principles of a human-centered medicine,^54^ others wrote also about the practitioners’ responsibility.^55^ And one of the few criticisms found on the concept, suggested that P4 “needs to be theoretically clarified and more widespread among - and dialogued with - health professionals of various specialties”^56^, and cannot be a medical practice only, but has to become an interdisciplinary practice.

The papers classified in the “specific iatrogenic examples” category addressed some specific population groups, for example pregnant women or mental health users. The authors generally criticized the medicalization of healthy people or physiological situations, empasizing the frequent iatrogenic results. We develop briefly here the examples of pregnancy and mental health. Pregnancy is a physiologic stage of life, however, health professionals usually handled it as a disease, especially the labour. Souza and Pileggi-Castro (2014) emphasized that the:

> Use of health technologies favors reduction in maternal morbidity and mortality, but hyper-medicalization – or the excessive and unnecessary use of health technology in care during pregnancy and childbirth – also represent risks for women, fetuses and newborns.^57^

Tesser and Norman (2015) reported the excess of episiotomy, cesarean section and routine use of ocytocine in Brasil. They also suggest PHC-centered P4 strategies, as the: elaboration of labor plans (personalized care itinerary), inclusion of trained family physicians to assist low-risk labour/childbirth and empowerment of the social movements which fight for “humanizing” it.^58^ In this regard, mental health represents a knowledge area whose epistemology, practice and results are frequently questioned.^59^ The selected papers of this topic point in the same direction: criticizing the treatment of healthy people, as well as the biological reductionist theories that are used to explain mental health problems, ruling out key elements like the path of life and other social dimensions. Another interesting example concerns the elderly people group. For multiple reasons, it constitutes a vulnerable group to iatrogenesis, because of usual: polymorbidity and polypharmacy, lower rate of metabolism (e. g.: renal clearance), cerebral and other functions (slower reflexion and understanding, difficulty of locomotion, hearing loss, etc.), shorter life expectancy, among others. In all these specific groups or situations, one frequent outlined strategy is “not to do”, using a “wait and see” approach, and sometimes even “desprescribing” suppressing prescriptions of medications and heavy treatments that are more harmful than beneficial to health. And to do so, the professionals need to have a good knowledge on EBM, so they can critically approach the available evidence, and adapt it to every individual situations. The other articles about odontology, pathology, chronic diseases developed similar reflections, reporting discrepancies between the excess of medical procedures, the cost-effectiveness and the non-maleficence principle.

Screening belongs to the second level of prevention and aims to detect asymptomatic diseases and treat them early. Consequently, screening procedures have to be realized, in the cases that the risks/benefits balance is unequivocally favorable to the patient. This differs completely from curative situations, where the patient has a disease and suffers; which allows a thinner margin between risks and benefits, because if we do not act, the clinical outcomes are much worse than if we do.^60^ Therefore, research on the harm of screening has to be enhanced and, maybe, connected with other type of iatrogenesis that can affect healthy people like primary prevention.

We will discuss further now the publications focused on the category “implementation and divulgation strategies of P4”. The authors proposed strategies intrinsically related to the drivers of medical overuse focusing on the main actors: physicians, patients, pharmaceutical industry, politicians as well as the means of communication between them. Improving the communication abilities of the physicians, for example, through a patient-centered approach^61^ was one of the most P4 suggested strategies. This approach involves: empathy, the construction of a trustworthy therapeutic relationship, exploring the illness, identifying the patient’s stress-factors and shared decision-making. All these communication skills evidenced effectiveness to improve mental health, reducing the overuse of health system and improving most of the symptoms. In this way, we understand that patient-centered medicine is one key element that could tackle medical overuse and reduce unnecessary medical procedures. Another significant and complementary perspective focuses particularly on the clinicians and invites them to enhance their critical skills to assess clinical guidelines, encouraging them, when possible, to use a watchful waiting approach and consequently avoiding overmedicalization. One of the selected papers^62^ mentioned the “Choosing Wisely” (CW) campaign, launched by the American Board of Internal Medicine in 2012^63^ that created and disclosed a list of five unnecessary medical procedures which had to be avoided and encouraged other medical societies to do the same. Moreover, this type of CW strategy showed evidences of cost-effectivess in multiple studies.^64^ Another suggested and pertinent strategy was the claim to reach a medicine without conflict of interests that biases most scientific studies and impacts strongly the guidelines. Consequently, it highly affects the clinical decision-making. To conclude this topic, we highlight an important aspect affecting the divulgation of the concept of P4. It is currently not well represented in the online search databases; and one of main causes is probably the fact that most of the databases does not classify it as a MeSH, or as a relevant keyword. Therefore, in order to boost significantly the promotion of P4 scientific identification and production, P4 needs to become a MeSH in the electronic databases.

### Strengths and weaknesses of the study

As we report in the introduction, the present study is the first bibliometric analysis and Descriptive Content Analysis on P4. While some bibliometric studies about adjacent topics like medicalization,^65^ overdiagnosis^66^ or “unnecessary procedures”^67^ does already exist, their objectives focused on some very specific clinical situations, such as: pregnancy or mammography screening. The last one focused on the impact of the Cochrane Reviews on clinical decision. For these reasons, we chose not to make any comparison between these publications and our study.

In addition, we employed a systematic methodology and used internationally acknowledged databases with minimal restriction criteria. Such sensitive approach positively supports the reliability of our results. However, in spite of our wide search methods, several key articles escaped our study and were not selected; some of them, probably because of the non-inclusion of P4 in the MeSH or keywords in the major databases, such as: one of Jamoulle (1986)^68^ and two articles of the reknowned Barbara Starfield, Gérvas and Heath.^69, 70^ Although these papers could have helped to promote the concept of P4, we chose not to include them for two reasons. First, because we would to highlight the existing impairment of P4 search in the selected databases. And second, their inclusion could invalidate our search method. Moreover, according to language criteria, another four articles had to be excluded because they were published in Serbo-Croat or Korean.

Furthermore, the selected papers had an heterogeneous number of authors and with a concern on a strong methodology, we decided to consider only the first authors data to perform the statistical results. This decision probably resulted in an overall reduction of the authors nationalities.

We knew that the journal ranking did not represent the most significant factor to assess the publications and Nature published about it in 2016:

> It effectively undervalues papers in disciplines that are slow-burning or have lower characteristic citation rates. Being an arithmetic mean, it gives disproportionate significance to a few very highly cited papers, and it falsely implies that papers with only a few citations are relatively unimportant.^71^

However, we decided to include the journal impact factor in our bibliometric analysis, because it is one of the few tools that can assess and, maybe, roughly explain the visibility – and not the quality – of some scientific studies in the world. Consequently, it could also help to understand and highlight journals and countries inequalities according to research output.

According to the method used in the selected articles, most of them were bibliographic studies. On the one hand, these types of studies provide interesting informations with conceptual or historical aspects that help to establish a robust background. But on the other hand, the studies were frequently non-systematized and aimed to support the concept without seeking a critical approach, thus weakening the content of those articles.

Several articles approached more than one category making it difficult to classify them under a single one. And with a concern on a global view on research output, we decided to classify each included paper under only one category according to the main article’s objective. So, as a qualitative classification, any bias according to the coder subjective decision could occur.

### Implication for practice

Quaternary Prevention represents a relevant concept for public health because it promotes the identification and prevention of potential iatrogenic situations and, consequently, can reduce overall iatrogenic mortality and health costs. In this respect, the authors suggest concrete implementation policies through: cross-sectoral collaboration (political, legal, social, educational and sanitary, among others), P4 teaching in the health graduation and post-graduation programs, improving the critical approach of clinical studies and guidelines, practical changes in the medical prescription and appropriate use of the communication skills by the physicians.

**WHAT IS ALREADY KNOWN ON THIS TOPIC**

- The concept of Quaternary Prevention (P4) has been designed initially at the end of the 1980s, when the Belgian family physician Jamoulle revised the prevention levels of Leavell and Clark according to the patient’s perception (illness) and the physician’s assessment (disease), integrating a public health approach into individual clinical practice.
- P4 is a recent theoretical concept for public health because it aims to tackle the outspread of unnecessary medical procedures and iatrogenic events; and consequently, can be able to lower global cost expenditures.

**WHAT THIS STUDY ADDS**

- Conceptual, geographical and linguistic aspects impacted the scientific production on P4, as well as the WONCA conferences and the model of healthcare systems in the authors’ country.
- The Brazilian were the first authors who most published (absolute number of publications) and the Uruguayan lead the production per capita.
- Most of the papers used a bibliographic research method and focused on the causes and examples of overmedicalization, just as strategies to implement P4.
- The quality and quantity of available studies is still limited and further investigations are recommended to assess the effective impact of P4 on public health.

### Implications for research

As we see above, Quaternary Prevention constitutes a critical approach on modern medicine and exposes most of its limitations. In this regard, P4 represents a counter-hegemonic concept and, consequently, research on it is clearly underfunded. While we observed a significant increase in the scientific production in 2015, the publications decreased again after this “golden year”. In this context, we consider that research to further the categories developed in this study is required. According to our analysis, three purposes have to be prioritized: the detection of overmedicalization situations (bibliographic investigative research), the identification of factors that lead to these situations and the development of P4 strategies (bibliographic research and qualitative studies) and testing the developed P4 strategies (quantitative studies).

For practical reasons and to provide effective results, we suggest that future research should focus on:

- Quantitative studies on situations of medical overuse, evaluating the iatrogenic outcomes of medical intervention (e.g.: primary and secondary prevention interventions) on previously healthy people;
- Studies assessing the impact of P4 strategies (e.g.: communication skills teaching, critical approach on EBM, disclosure of the physician’s conflict of interests) on clinical outcomes (mortality, iatrogenic events, cost-effectiveness, patient and physician’s perceptions), and;
- Studies and interventions of health promotion (e.g.: physical activity, nutritional, stress management, social inclusion measures) and/or health education (e.g.: population education about P4), without medical interventions and analyzing the outcomes (e.g.: mortality, cost-effectiveness, patient and physician’s perceptions);

In addition, the total number of articles remains low if compared to other search terms like “medical overuse” which have up to 100 times more results than “quaternary prevention” in Pubmed. We understand that a linguistic and conceptual gap could explain this consistent difference and consider that some strategies could be able to transcend it, involving linguistic, conceptual, information and communication technologies dimensions, respectively:

- To insist on the English language to promote P4 publications worldwide;
- To disclose the concept and clarify possible misunderstanding regarding the initial and actual acknowledged definition of the concept;
- To come closer to other adjacent and more “famous” concepts, such as “medical overuse”, “overdiagnosis” or even “overtreatment” and put them in the abstracts and as keywords;
- To publish more often in Q1 or Q2 reviews and, when possible, addressing quantitative studies that support P4 (e. g.: cost-effectiveness and mortality reduction);
- To claim the inclusion of P4 as a MeSH in the international databases.

## Conclusion

The present bibliometric and content analysis highlighted key elements that affected the research output on P4, such as: conceptual, geographical, linguistic, WONCA conferences and model of healthcare systems in the authors’ country. Most of the included papers used a bibliographic research method and approached quite equally the causes, the examples of overmedicalization and the implementation strategies. Quaternary prevention represents an ethical combat towards fairness and equity of access to health services. The concept and its respective scientific output are relevant for public health, however we consider that there is still a limited quality and quantity of available studies on the topic and further studies are recommended, especially in order to assess its effectiveness to prevent the outbreak of iatrogenic events and to lower global health expenditure.

## Data Availability

Not applicable.

## Notes

### Competing Interest Statement

The authors have declared no competing interest.

### Funding Statement

No external funding was received.

### Author Declarations

All relevant ethical guidelines have been followed and any necessary IRB and/or ethics committee approvals have been obtained.

Any clinical trials involved have been registered with an ICMJE-approved registry such as ClinicalTrials.gov and the trial ID is included in the manuscript.

